# Understanding the atopic dermatitis-psoriasis phenotypic switch through a mechanistic epidemiology approach

**DOI:** 10.1101/2025.05.25.25328309

**Authors:** Kerry Yang, Alexandra Mircescu, Deborah Okusanya, Samiha Mohsen, Danlin Zeng, Sonia Czyz, Isabelle Vallerand, Giovanni Damiani, Christopher G. Bunick, Fatemeh Jafarian

## Abstract

Atopic Dermatitis (AD) and psoriasis (PsO) are two frequent dermatologic conditions that may co-occur in a cluster of patients, yet current understanding of how these two conditions relate to one-another remains poorly understood. One way to better understand their relationship is through a process called phenotypic switching, where AD and PsO can turn into one another. We utilized a pharmacovigilance-based epidemiological approach to better understand this phenomenon. By generating adverse event-related disproportionality signals for various therapies and therapeutic classes used in AD and PsO, several potential mechanisms for the AD-PsO phenotypic switch were uncovered. This includes mechanisms involving T_H_2 and T_H_22 repolarization, T_H_17 and T_H_22 repolarization, and immune shifting between T_H_1, T_H_17, and T_H_2 cells. Clinically and immunologically related conditions were also analyzed to gain a clearer understanding of the specificity of the switch from PsO to an eczematous phenotype. Together, these findings provide mechanistic insight into the underpinnings behind the AD-PsO phenotypic switch through a novel approach, adding evidence to the fluid nature of immune phenotypes in common medical conditions.

**One Sentence Summary:** The atopic dermatitis-psoriasis phenotypic switch creates an overlap phenotype that is likely T_H_22-driven.

## INTRODUCTION

Atopic Dermatitis (AD) and psoriasis (PsO) are two of the most common immune-mediated skin conditions worldwide. The global prevalence of AD is estimated to be around 2-4%(*1*), with a lifetime prevalence of up to 20%(*2*). PsO has a global prevalence ∼2-3%. AD presents clinically as intensely pruritic, erythematous, scaly lesions that preferentially affect flexural areas like the antecubital and popliteal fossae. AD typically starts in early childhood, often undergoing a relapsing-remitting pattern, and is associated in the atopic triad with asthma and allergic rhinitis. AD is primarily driven by the type 2 (T_H_2) immune response, although the type 1 (T_H_1), type 17 (T_H_17), and type 22 (T_H_22) responses are involved as well(*2*). In contrast, PsO presents with well-demarcated, erythematous plaques containing thick, silvery-white scales that often involves extensor surfaces, and specialized sites like the nails, scalp, genitalia, and intertriginous body folds. PsO is primarily driven by the T_H_17 and T_H_1 immune responses(*3*). Both conditions can significantly impact quality of life,(*4–6*) alongside strong associations with various atopic, autoimmune, cardiovascular, metabolic, and psychiatric comorbidities(*7*, *8*).

AD is associated with significant pruritus, xerosis, and often fluctuates in course(*2*, *9*). The adaptive T cell response during the acute phase of AD predominantly involves the T_H_2 immune response, with interleukin (IL)-4, IL-5, IL-13, and IL-31 all playing an important role in disrupting the skin barrier and inducing itch, contributing to the presentation of the disease(*10*). T_H_22 cells and IL-22 also play a sizable role(*11*). In chronic AD, IFN-γ is upregulated, and this can lead to down-regulation of ceramide synthase perpetuating the inflammation-barrier disruption cycle(*12*). Along with mainstay treatments, targeted therapies approved for the management of AD include the IL-4Rα inhibitor dupilumab(*13*), which reduces IL-4/13 signaling, and the IL-13 inhibitors tralokinumab(*14*) and lebrikizumab(*15*). More recently, an IL-31R inhibitor nemolizumab(*16*) was approved for use in AD(*17*). Oral Janus Kinase (JAK) inhibitors, abrocitinib and upadacitinib(*18*), are approved in various countries for AD treatment. The JAK-STAT signaling pathway plays an important role in activating downstream gene transcription for various cytokines involved in inflammation and pruritus. Inhibiting this pathway effectively manages immune-mediated conditions such as AD.

PsO pathogenesis is dominated by T_H_17 cells, but T_H_1 and T_H_22 also play a significant role(*3*). T_H_1 is activated by IL-12, while T_H_17 and T_H_22 are activated by IL-23. These activated cell types produce proinflammatory cytokines like IFN-γ, TNF-α, and IL-22, promoting inflammation and epidermal hyperproliferation(*3*). T_H_17 also produces several subtypes of IL-17 (ie, IL-17A, IL-17F), which positively feedback to T_H_17 to sustain the disease(*19*). Current biologics approved for moderate-to-severe PsO include TNF-α inhibitors adalimumab, certolizumab pegol, etanercept, golimumab, and infliximab, IL-17 inhibitors secukinumab, ixekizumab, brodalumab, and bimekizumab, IL-23 inhibitors guselkumab, tildrakizumab, and risankizumab, and the IL-12/23 inhibitor ustekinumab(*3*). Oral allosteric TYK2 inhibitors like deucravacitinib are also approved for use in treating PsO(*20*), as well as phosphodiesterase-4 inhibitors such as apremilast(*21*).

AD-related barrier dysfunction can lead to more frequent self-stranded DNA recognition and thus lead to the development of psoriasis(*22*). At the same time, the systemic inflammation underlying psoriasis can trigger dysfunctional maturation of keratinocytes, leading to AD in predisposed individuals.(*23*)

As AD and PsO research evolves, overlapping immunological pathogenesis and disease characteristics have emerged, suggesting these two conditions potentially exist on a spectrum of disease rather than in isolation(*11*, *24–26*). Along with cases where both conditions exist concurrently(*27*) or when an overlap condition develops showing clinical features of both(*28*), an interesting phenomenon that occurs is when a patient with AD phenotypically switches into PsO, or vice versa, with or without treatment(*29*, *30*). Also called “paradoxical reactions”, reports exist of PsO developing in AD patients treated with dupilumab(*31–36*) rather than other humoral autoimmune diseases(*37*), along with AD developing in patients with PsO(*38*, *39*), including those treated with TNF-α(*40–42*), IL-17 (*36*, *43–47*), IL-12/23(*48–50*) and IL-23 inhibitors(*51*, *52*). AD is also known to form eczematous phenotypes after exposure to dupilumab(*53*, *54*)

Phenotypic switching is a commonly occurring phenomenon. It has been observed by various cell populations when adapting to changes in the environment(*55*), One such example is the transition of tumor cells from a drug-sensitive to a drug-resistant phenotype in the treatment of various cancers(*56*, *57*). It can also occur with immune cells, as exemplified by the AD-PsO paradoxical reaction.

Even with advances in understanding the immunological overlap between PsO and AD, the AD-PsO phenotypic switch remains poorly understood. IL-22, produced by T_H_17 and T_H_22 cells, is implicated in both skin conditions, highlighting an immunological linkage and potential common mechanism between the two diseases (*11*).

One proposed theory for the paradoxical emergence of AD after PsO biologics is that inhibiting T_H_1/ T_H_17 cytokines induces a shift to a T_H_2-dominated response, resulting in an eczematous phenotype(*30*). Phenotypic switching to T_H_2-mediated conditions has been observed when immunomodulation suppresses T_H_1/ T_H_17 responses(*38*). For example, TNF-α inhibitors are associated with a downregulation of the T_H_1/ T_H_17 immune axis, and upregulation of the T_H_2 immune axis(*58*, *59*). These inhibitors are associated with the development of psoriasiform dermatitis, which is an intermediary phenotype that tends to have the clinical characteristics of

## RESULTS

PsO, but is histologically similar to AD(*60*, *61*). To better understand the relationship between biologics and the AD-PsO phenotypic switch, we utilized a pharmacovigilance-based disproportionality approach using data from the FDA Adverse Events Reporting System (FAERS)(*62*).

### Biologics used in atopic dermatitis create a transitory phenotype between atopic dermatitis and psoriasis

AD is driven by a T_H_2 response and PsO is driven by a T_H_17 response, with sizable contribution from T_H_1 (Fig. 1). The biologics approved for the treatment of AD include the IL-4/13 inhibitor dupilumab and the IL-13 inhibitor. A pharmacovigilance-based disproportionality analysis on FAERS data was run to see if signals emerged for PsO and its subtypes in relation to dupilumab or tralokinumab(*62*) (**Methods**).

**Figure 1.**
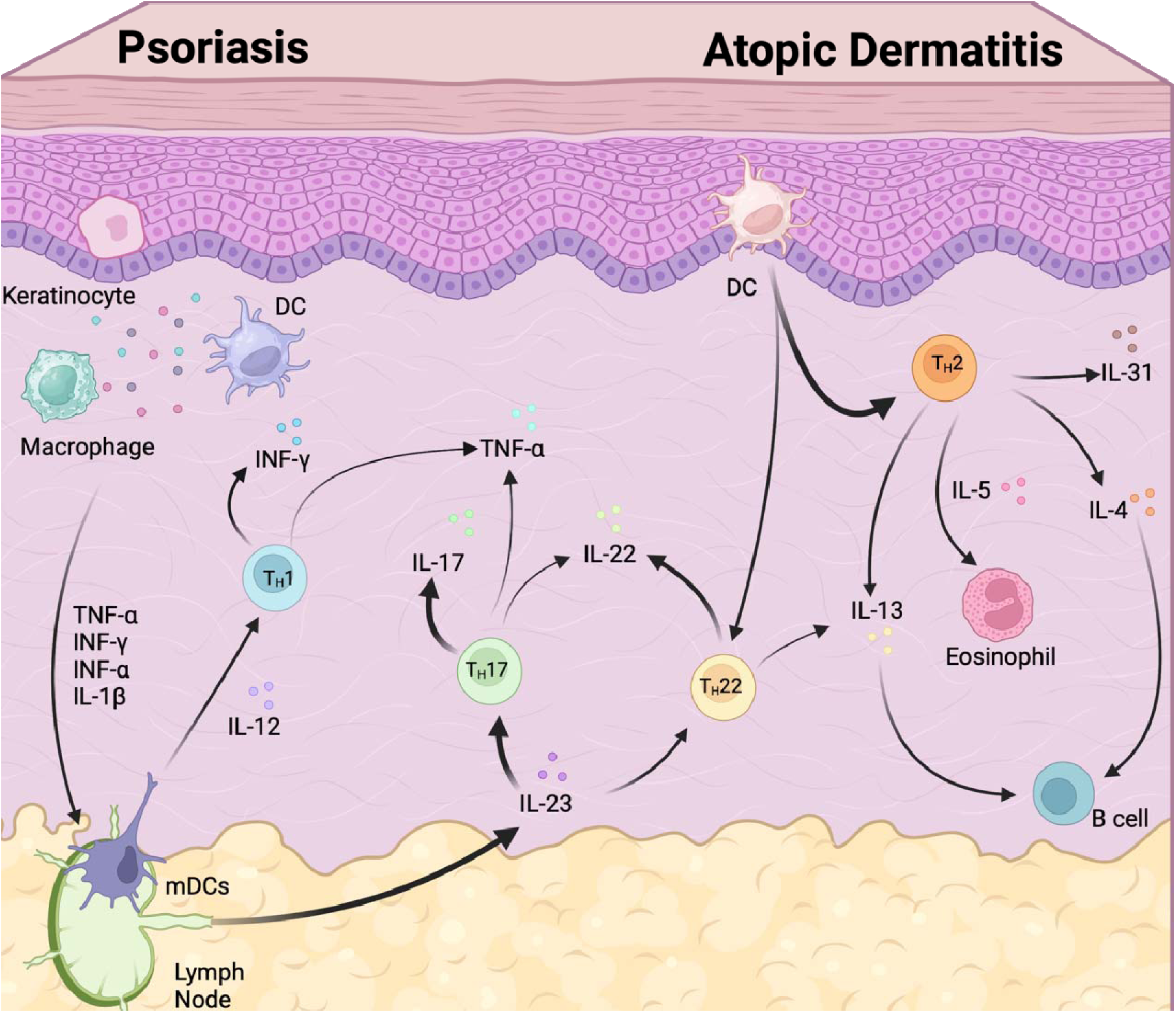
Pathogenesis of Atopic Dermatitis and Psoriasis. Illustration of the immunopathogenesis of atopic dermatitis (AD) and psoriasis (PsO). In AD, impaired skin barrier allows allergen penetration, initiating a T-Helper (T_H_) 2-dominant inflammatory response. Key cytokines (IL-4, IL-5, IL-13, IL-22) promote IgE production, eosinophil recruitment, and barrier dysfunction. Psoriasis is triggered by environmental stressors. These stressors activate innate immunity and leading to T_H_1, T_H_17, and T_H_22 responses via cytokines such as TNF-α, IL-12, and IL-23. Phenotypic overlap occurs through shared immune dysregulation.

Among PsO-related adverse events, psoriasiform dermatitis was the only meaningful signal to emerge for dupilumab (ROR [95% CI] = 2.64 [2.07, 3.36]) when no treatment indication was specified. The signal for PsO was 0.57 [0.54, 0.60]. When the indication for use was set to AD, eczema, or dermatitis, the ROR for psoriasiform dermatitis changed to 3.23 [2.37, 4.39] for dupilumab, while the PsO signal was 0.56 [0.52, 0.61] (Fig. 2A). The signal strengthened for psoriasiform dermatitis while the PsO signal remained stable (Fig. 2B). No signals related to tralokinumab met the inclusion criteria (Fig. 2C).

**Figure 2.**
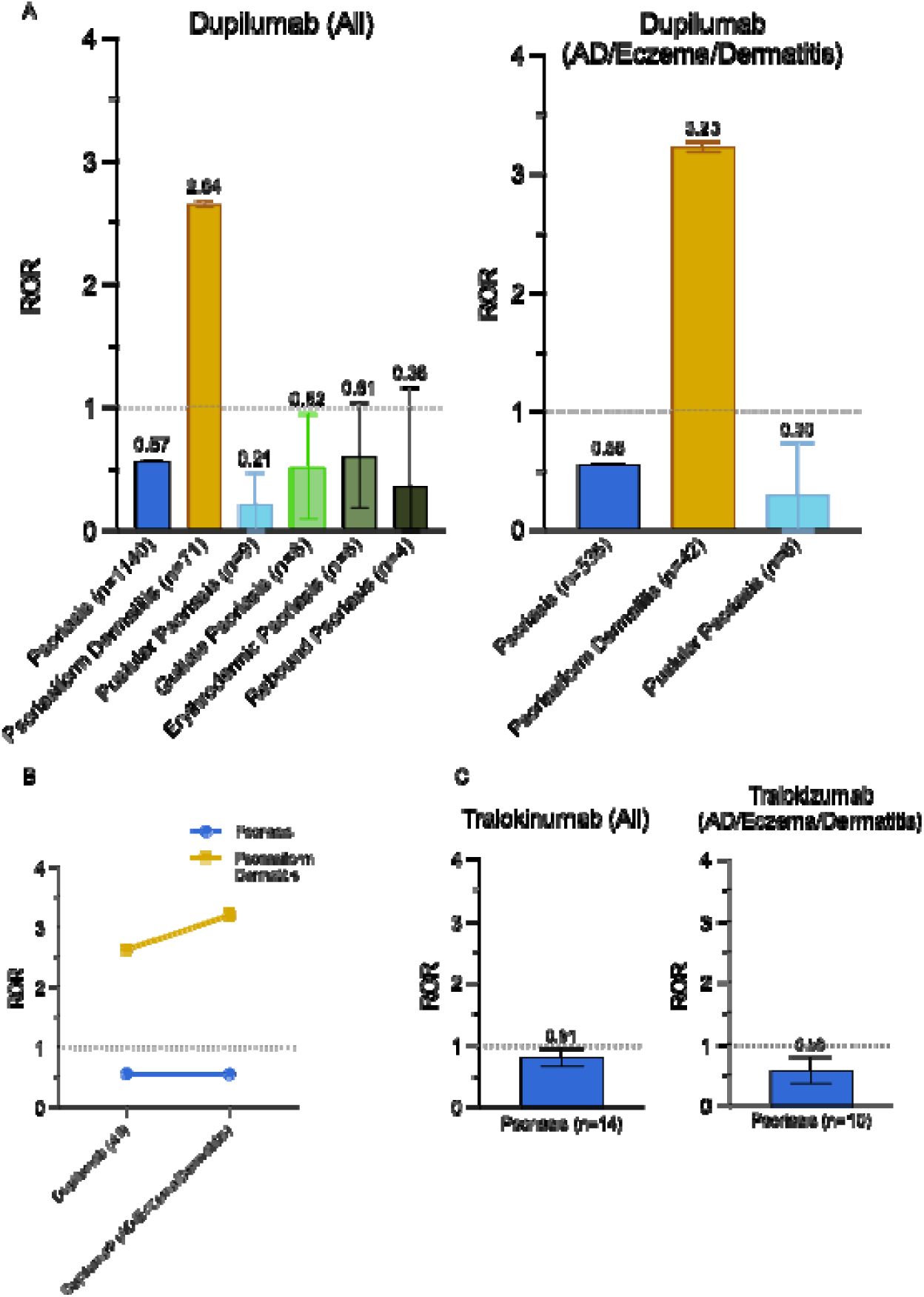
Psoriasis-Related Adverse Events with Biologics for Atopic Dermatitis. (**A**) Reporting odds ratios (RORs) for psoriasis-related adverse events with dupilumab, across all indications and restricted to AD-related indications. (**B**) Change in ROR signal strength for psoriasis and psoriasiform dermatitis with narrowed indications. (**C**) Psoriasis-related RORs for tralokinumab across all indications and for AD-specific use. Error bars represent confidence intervals.

### No adverse event signals for other T_H_1 and T_H_17-driven immune-mediated conditions were found

Other common diseases involving the T_H_1 and T_H_17 pathways include inflammatory bowel disease (IBD), the inflammatory arthropathies such as rheumatoid arthritis and ankylosing spondylitis, systemic lupus erythematosus, and multiple sclerosis. These conditions and their related terms on FAERS were analyzed to see if adverse event signals emerged for dupilumab and tralokinumab (**Fig. S1**). No signals were found, both in general (**Fig. S1A**) and with AD, eczema, or dermatitis as an indication (**Fig. S1B**), suggesting the T_H_1/ T_H_17 inflammatory cascades do not play major roles in the AD to PsO phenotypic switch.

### Average eczematous dermatitis RORs increased with PsO but not with IBD

Eczematous dermatitis is a term used to classify a range of inflammatory skin conditions that share similar clinical and histological characteristics(*63*). These include conditions such as atopic, contact, dyshidrotic, neuro, nummular, seborrheic and stasis dermatitis. Although there are immunological differences, they have clinical and histological overlap. The RORs for various eczematous dermatoses were given equal weighting and averaged together to reduce noise and reporting bias that is often found within FAERS. The average RORs for each class or medication in general was compared to the average RORs when the indication was set to PsO, Crohn’s disease (CD), or ulcerative colitis (UC) (Fig. 3).

**Figure 3.**
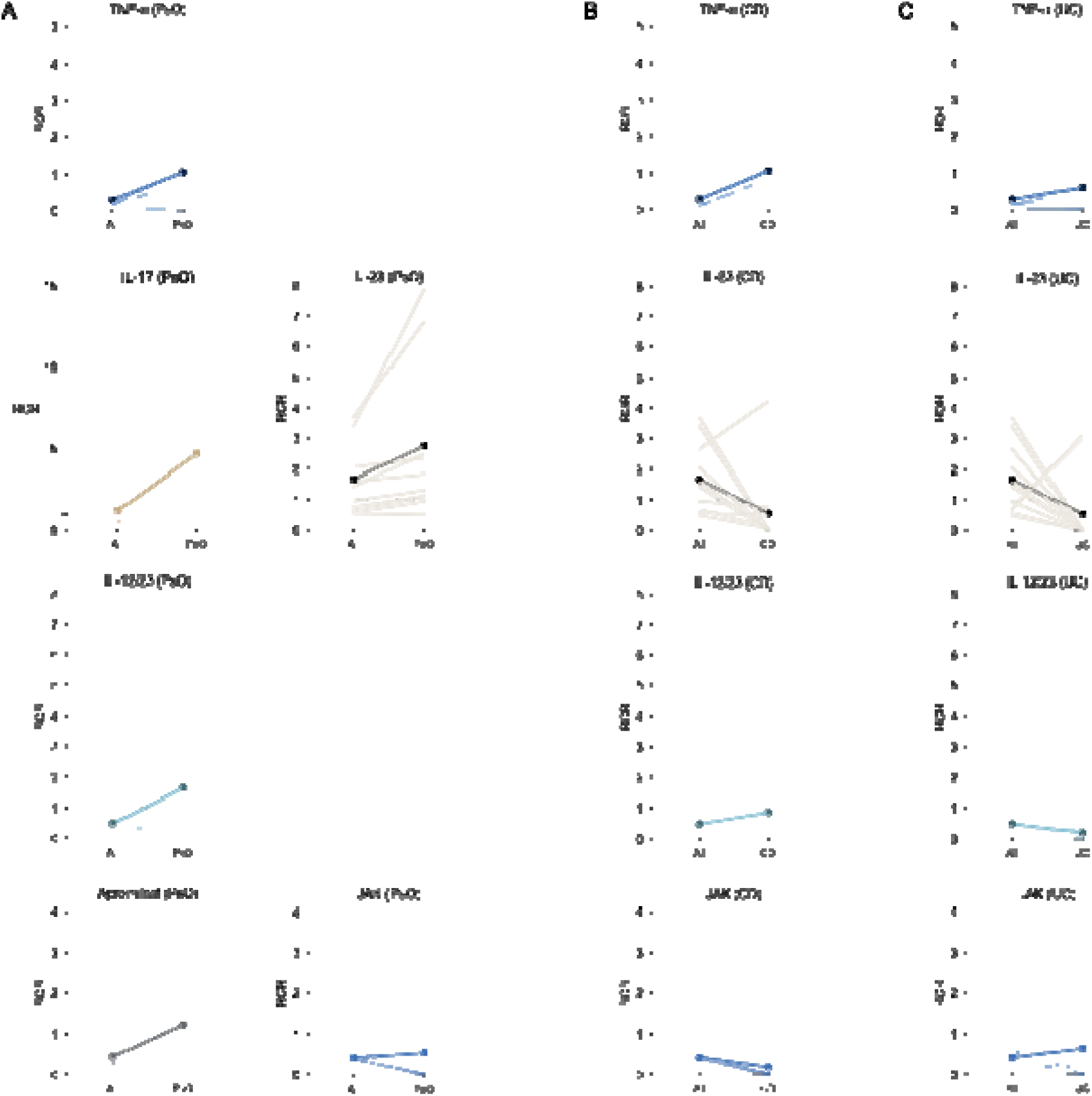
Reporting Odds Ratios for Eczematous Dermatitis Across Conditions. Change in average RORs for eczematous dermatoses with various therapies when the treatment indication was specified as: (**A**) Psoriasis (PsO), (**B**) Crohn’s disease (CD), (**c**) Ulcerative colitis (UC). Included adverse events span MedDRA-coded eczematous conditions (e.g., atopic, allergic, seborrheic, dyshidrotic dermatitis). Equal weighting was applied to minimize reporting bias. Events with fewer four occurrences were excluded.

IBD, namely CD and UC, shares similar pathophysiological features with PsO, which is why they have overlapping treatment options(*64*). Some shared medications, such as TNF-α inhibitors, cause psoriasiform skin lesions when treating IBD(*65*). To explore whether the phenotypic switch to AD is unique to PsO, the changes in signal strength for common medications and medication classes were compared between PsO and IBD.

This analysis looked at the TNF-α inhibitors adalimumab, certolizumab pegol, etanercept, golimumab, and infliximab, the IL-17 inhibitors bimekizumab, brodalumab, ixekizumab, and secukinumab, the IL-23 inhibitors guselkumab, risankizumab, and tildrakizumab, the IL-12/23 inhibitor ustekinumab, the phosphodiesterase-4 inhibitor apremilast for psoriasis. The JAK-inhibitors deucravacitinib, ruxolitinib, tofacitinib, and upadacitinib were also included as a control group for psoriasis. The IBD analysis included the TNF-α inhibitors adalimumab, etanercept, golimumab, and infliximab, the IL-23 inhibitors guselkumab, mirikizumab, risankizumab, and tildrakizumab, and the IL-12/23 inhibitor ustekinumab and the JAK-inhibitors tofacitinib and upadacitinib. Other analyses included the α4β7 integrin inhibitor vedolizumab.

Looking at the change in ROR when limiting the indication from all conditions to PsO, there is a noticeable increase in all medications and medication classes, indicating the PsO to AD switch is not only due to the medications themselves, but due to the relationship between PsO and AD (Fig. 3A). When looking at CD and UC, there is no clear trend in the formation of eczematous dermatoses, and with many RORs remaining or dropping below 1 (Fig. 3**, B and C**), suggesting the paradoxical reaction is specific to PsO and not IBD. IBD and PsO likely form different skin lesions even when using similar immunomodulating therapies, although there might be a shared pathway with TNF-α inhibitors. The PsO to AD phenotypic switching effect is also quite strong given the changes in signal strength, especially with IL-17 inhibitors (Fig. 3A).

### PsO therapies create a transitory phenotype between AD and PsO

An analysis was performed for the medications in general (Fig. 4A) and with PsO as an indication (Fig. 4B). When looking at adverse events related to the psoriasis-related immunomodulatory therapies in general, signals did not emerge for eczema, dermatitis, and atopic dermatitis. Dermatitis psoriasiform signals emerged for TNF-α (1.51 [1.35, 1.69]), IL-17 (3.89 [3.30, 4.58]), and IL-12/23 (1.79 [1.33, 2.41]) inhibitors. Signals did not emerge for apremilast (0.95 [0.62, 1.45]) nor JAK-inhibitors (0.61 [0.54, 0.68]). The signal for IL-23 inhibitors was nonsignificant given the inclusion criteria (1.44 [0.82, 1.45], χ^2^_Yates_=1.19) (Fig. 4A).

**Figure 4.**
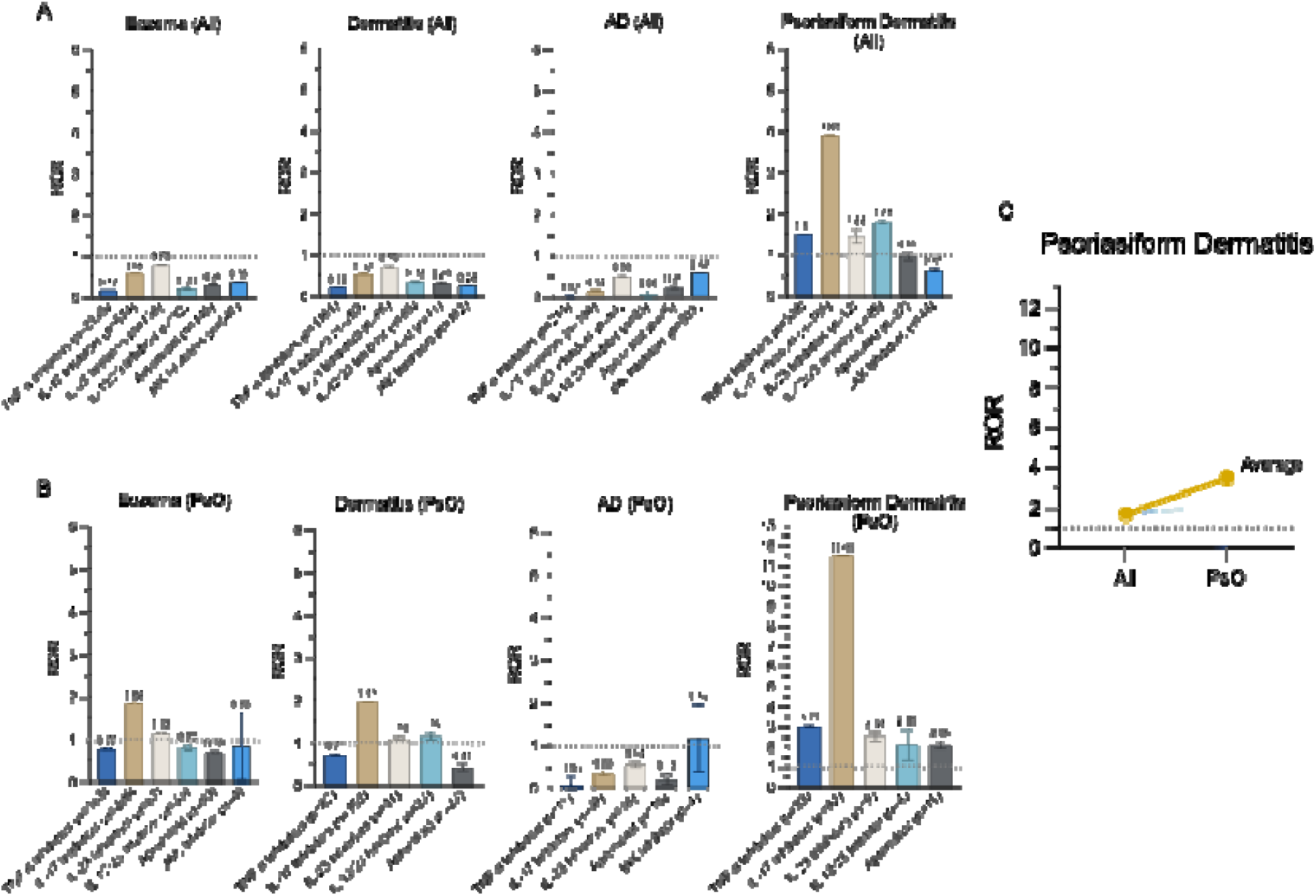
Atopic Dermatitis-Related Adverse Events from Psoriasis Therapies. (A) RORs for eczema, dermatitis, AD, and psoriasiform dermatitis with immunomodulatory therapies across all indications. (B) Same outcomes when therapies were used specifically for PsO. (C) Change in psoriasiform dermatitis RORs with indication narrowed from all conditions to PsO.

When looking at cases with PsO as an indication, IL-17 inhibitors had signals emerge for eczema (1.84 [1.64, 2.06]) and dermatitis (1.95 [1.66, 2.29]), while the others did not. There were no significant signals for atopic dermatitis. Psoriasiform dermatitis signals increased for TNF-α (3.01 [2.13, 4.26]) and IL-17 (11.46 [9.16, 14.34]) inhibitors, while signals emerged for IL-23 inhibitors (2.49 [1.29, 4.80]) and apremilast (2.04 [1.13, 3.71]). There were no psoriasiform dermatitis cases with JAK-inhibitors. The ROR for the IL-12/23 inhibitor ustekinumab was higher (2.06) when PsO was an indication compared to ustekinumab in general, but it did not meet the signal inclusion criteria ([0.77, 5.50], χ^2^_Yates_ = 1.25) (Fig. 4B).

Each immunomodulatory therapy has their own unique adverse event signal profile. The overall increased signal strength for all therapies when PsO was set as an indication, compared to the RORs for the immunomodulatory therapies in general, points towards the paradoxical reaction being related to PsO itself and not only due to the medications. The signals were stronger for psoriasiform dermatitis compared to eczema, dermatitis, and AD (Fig. 4**, A and B**), and there was a net increase in psoriasiform dermatitis signal strength when treating PsO (Fig. 4C).

### Other eczematous adverse event signals emerged for PsO-related therapies

When looking at the analyzed therapies taking all indications into account, eczematous dermatitis signals emerged for IL-17 (seborrhoeic dermatitis 1.86 [1.44, 2.41], dyshidrotic eczema 3.11 [2.26, 4.29], and stasis dermatitis 1.77 [1.15, 2.73]) and IL-23 inhibitors (seborrhoeic dermatitis 2.65 [1.62, 4.35] and hand dermatitis 3.42 [1.69, 6.89]). Generalized exfoliative dermatitis, a drug hypersensitivity reaction, was present for IL-23 inhibitors (4.10 [3.23, 5.21]) and IL-12/23 (1.32 [1.04, 1.68]). Dermatoses related to infections, such as infected dermatitis (IL-17 inhibitors 1.87 [1.20, 2.91] and apremilast 2.53 [1.48, 4.32]) as well as eczema herpeticum (4.20 [3.11, 5.68]) emerged as well (**Fig. S2A**).

When PsO was an indication, significantly more signals emerged for dermatoses that fall under the eczematous dermatitis category, including contact dermatitis (TNF-α inhibitors 1.33 [1.07, 1.67]), seborrheic dermatitis (IL-17 inhibitors 7.28 [5.28, 10.04], IL-23 inhibitors 3.81 [2.04, 7.10], and apremilast 2.30 [1.19, 4.43]), stasis dermatitis (TNF-α inhibitors 3.03 [1.56, 5.88], IL-17 inhibitors 8.62 [5.29, 14.04], IL-12/23 inhibitor 7.67 [2.86, 20.55], and apremilast 2.75 [1.03, 7.38]), and dyshidrotic eczema (IL-17 inhibitors 12.74 [8.67, 18.72]), among others (**Fig. S2B**). Looking at overall trends, IL-17 inhibitors seem to have more signals, and stronger signals overall, for eczematous dermatoses, adding further credence to the role of T_H_17 in the PsO to AD phenotypic switch.

More exfoliative dermatitis (TNF-α 2.20 [1.57, 3.07] and IL-12/23 8.30 [5.50, 12.52] inhibitors) and generalized exfoliative dermatitis (TNF-α 5.23 [4.18, 6.54], IL-17 4.36 [3.42, 5.56], and IL-12/23 3.51 [2.08, 5.94] inhibitors) signals emerged when PsO was set as an indication. Adverse event signals associated with infections that have dermatitis or eczema as keywords include infected dermatitis (TNF-α 2.59 [1.22, 5.48], IL-17 inhibitors 7.79 [4.55, 13.31], IL-23 inhibitors 4.53 [1.69, 12.15], and apremilast 3.04 [1.13, 8.16]) and acarodermatitis (TNF-α 2.32 [1.39, 3.88], IL-23 inhibitors 3.80 [1.89, 7.62], and apremilast 7.09 [3.53, 14.24]) increased as well. The only signal that emerged for JAK inhibitors was dermatitis acneiform 12.34 [6.16, 24.70], adding support to the existence of JAK-induced acne (**Fig. S2B**).

### Signals emerged for other allergic adverse events in PsO-related therapies

Allergic diseases are conditions that are characterized by aberrant immune responses to normally innocuous antigens. Conditions include the atopic triad of AD, asthma, and allergic rhinitis, along with food allergies and anaphylaxis. Although many elements play key roles in their pathogenesis(*66*), allergic conditions are driven by T_H_2 cells and their cytokines(*67*). Common cytokines produced by T_H_2 cells include IL-4, which induces B cell isotype switching to produce immunoglobulin E (IgE), IL-5, which promotes eosinophil differentiation, migration, and survival, and IL-13, which induces mucus hypersecretion and fibrosis(*67*, *68*). To examine if inhibition of T_H_1/ T_H_17 cytokines involved in psoriasis(*69*) would increase T_H_2 activity, a disproportionality analysis was run on several conditions containing allergy-related keywords (**Methods**) (Fig. 5). Allergic edema (3.90 [2.37, 6.42]), allergic pruritus, (2.02 [1.19, 3.43]), allergic cough (2.69 [1.55, 4.66]), allergic sinusitis (2.38 [1.31, 4.32]), and allergic respiratory symptom (3.26 [1.45, 7.34]) emerged as signals for TNF-α inhibitors, while multiple allergies (2.20 [1.54, 3.13]), allergic pruritus (4.44 [2,38, 8.29]), and allergic edema (5.17 [2.45, 10.91]) were positive signals for IL-23 inhibitors. Rhinitis (2.58 [2.05, 3.25], 2.72 [1.75, 4.21]) emerged for IL-17 and IL-12/23 inhibitors, respectively, and multiple allergies was 3.95 [3.17, 4.91] for apremilast (Fig. 5A). Taken together, these results suggest that downregulating the T_H_1/ T_H_17 immune axis may be linked with a corresponding increase of the T_H_2 axis, leading to increased allergy-related conditions. This is especially true of TNF-α inhibitors.

**Figure 5.**
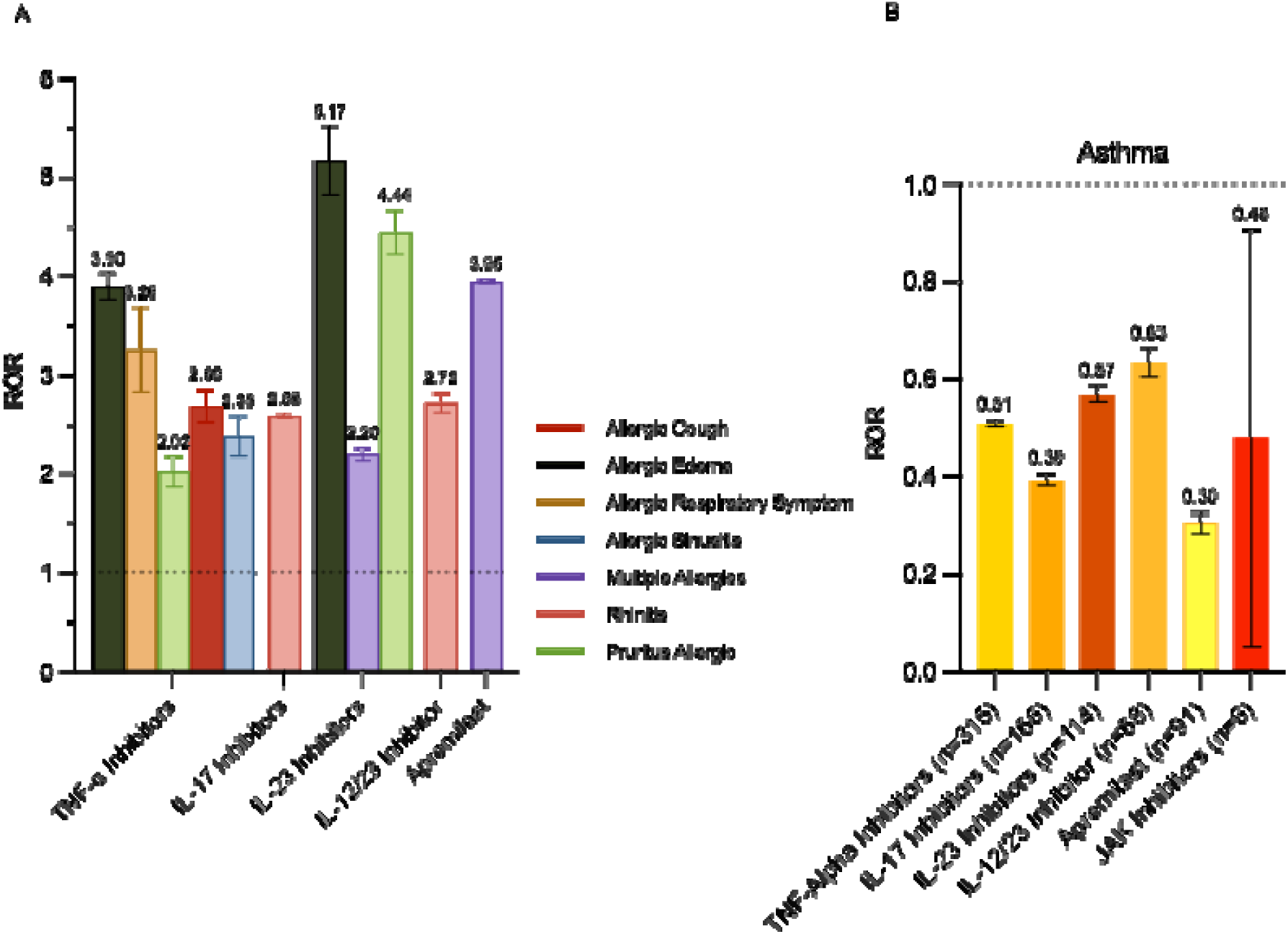
Allergy-Related Adverse Events in Psoriasis Treatment. (A) Allergy-Related adverse events associated with PsO therapies that met inclusion criteria. (B) Asthma-specific RORs across medications when the indication was PsO.

### A signal for asthma did not emerge in PsO-related therapies

Asthma is a common chronic respiratory disease and part of the atopic triad with AD and allergic rhinitis. It is immunologically quite heterogeneous with multiple immune pathways and endotypes involved(*70*), which could explain the heterogeneity in treatment outcomes and adverse events with biologics targeting T_H_2-associated cytokines(*71*, *72*). It is postulated that asthma likely involves T_H_1, T_H_2, and T_H_17 cells, with the T_H_2 asthma endotype being associated with elevated eosinophil levels, and the T_H_17 endotype associated with neutrophilic asthma(*70*). T_H_22 also plays a significant role in asthma pathogenesis, likely in an anti-inflammatory manner(*73*). A disproportionality analysis was done for asthma as an adverse event with PsO-related medications (Fig. 5B). No asthma signals emerged (TNF-α, 0.51 [0.46, 0.57], IL-17 inhibitors, 0.39 [0.34, 0.46], IL-12/23 inhibitor, 0.63 [0.50, 0.80], IL-23 inhibitors, 0.63 [0.50, 0.80], apremilast, 0.30 [0.25, 0.37], and JAK inhibitors, 0.48 [0.21, 1.07]), suggesting PsO does not phenotypically switch into all atopic conditions.

## DISCUSSION

AD is primarily driven by the T_H_2 pathway and its associated cytokines, which includes IL-4, IL-5, and IL-13, and in some endotypes by T_H_22, T_H_1 and T_H_17 pathways. PsO is driven by the T_H_1 and T_H_17 pathways, along with the associated cytokines IL-12, IL-23, IL-17, and IL-22.

Importantly, both conditions have a shared pathway through T_H_22. Our results suggest the AD-PsO phenotypic switch leads to the creation of an intermediary condition with shared features, likely through common pathways linking the anti-IL-4/13 driven switch in AD and the anti-IL-17 driven switch in PsO. (Fig. 6). AD likely develops a psoriasiform phenotype when treated with dupilumab through the inhibition of the T_H_2 inflammatory cascade. This causes a shift towards an increased T_H_22 response, thus creating a psoriasiform dermatitis pattern (Fig. 6A)(*53*).

**Figure 6.**
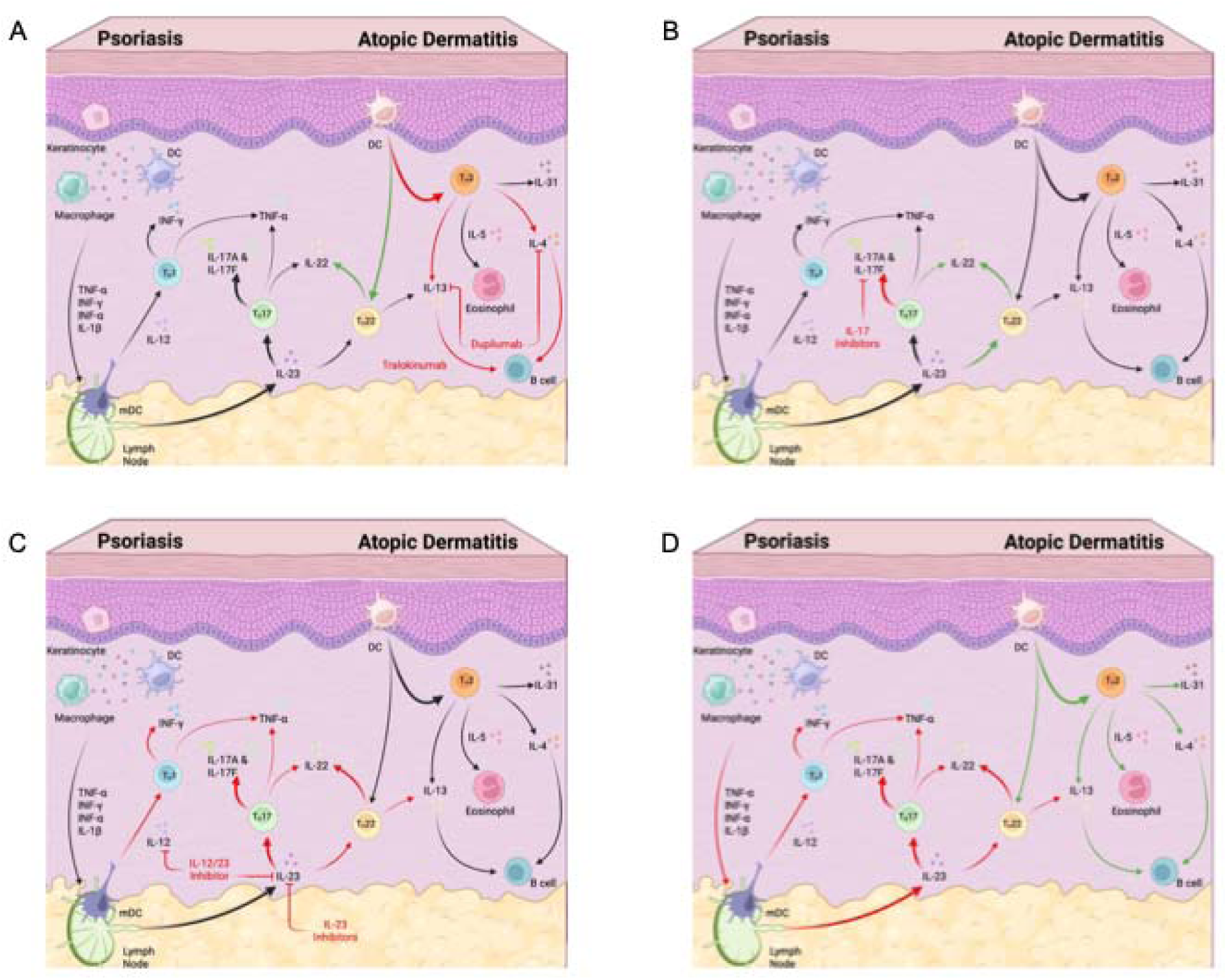
Mechanisms Behind the Atopic Dermatitis-Psoriasis Phenotypic Switch. Proposed immune mechanisms explaining paradoxical switching between AD and PsO: (A) IL-4/13 inhibition in AD therapy upregulates T_H_22. (B) IL-17 blockade increases T_H_22 responses. (C) IL-23 inhibition (with or without IL-12) prevents switching via T_H_17/T_H_22 pathway. (D) Downregulation of T_H_1/T_H_17 permits T_H_2 upregulation.

The reason why IL-17 inhibition has a much stronger PsO to AD switching effect than other classes of biologics is likely due to the differentiation of T cells into T_H_17 and T_H_22 (Fig. 6B). TNF-α and IL-23 are known to support both T_H_17 and T_H_22 production and differentiation(*74*– *76*), indicating the T-helper cell types have a shared upstream pathway. T_H_22 also plays a significant role in the acute phase of AD(*77*). Inhibiting the downstream effects of the T_H_17-mediated inflammatory cascade likely increases T_H_22 polarization, which would explain the unexpected strength of psoriasiform dermatitis with IL-17 inhibition compared to the rest.

Since TNF-α, IL-23, and IL-12/23 inhibitors work upstream of T_H_17 and T_H_22 cells, they do not change the dynamics of the T_H_17/T_H_22 axis like IL-17 inhibitors (Fig. 6C). Instead, given the signals related to the formation of T_H_2-related conditions (Fig. 5), downregulation of the T_H_1/T_H_17 axis with increased activity of the T_H_2 immune axis would be the likely mechanism for TNF-α, IL-12/23, and IL-23 inhibitors (**Fig 6D**)(*58*, *59*). This mechanism appears to be much weaker than the shift in T_H_17 and T_H_22 polarizations with anti-IL-17 biologics.

This study provides pathogenetic insights through a pharmacovigilance-based disproportionality analysis using FAERS, utilizing “mechanistic epidemiology” to understand the AD-PsO phenotypic switch. Here, our results show that dupilumab used in AD and PsO can create an intermediary histological and immunological phenotype in the AD-PsO phenotypic switch. For the AD to PsO phenotypic switch, the postulated mechanism is downregulation of the T_H_2 cytokine cascade causes upregulation of T_H_22 cells, and thus a more psoriasiform phenotype(*53*), without as much involvement from T_H_1 and T_H_17 cells. Conjunctivitis, blepharitis, and rosacea can arise from dupilumab use as well, and it seems that lebrikizumab might be a better choice if these adverse events arise.

For the PsO to AD phenotypic switch, IL-17 inhibitors are much more likely to cause the paradoxical reaction compared to other therapies used in PsO. Given our current understanding on PsO pathogenesis, the most likely mechanism to explain this unexpected finding is a change in the T_H_17/T_H_22 immune polarization. Inhibition of the downstream immune response of T_H_17 causes increased T_H_22 polarization, leading to a disease phenotype that shares characteristics with AD(*73*). Our findings also suggest that downregulation of immune pathways more strongly associated with PsO, such as T_H_1 and T_H_17, can lead to potential upregulation of other immune pathways, such as the T_H_2 pathway and its associated cytokines, although this is a weaker effect than shifting the polarization between T_H_17 and T_H_22 cell subtypes.

Providers are recommended to be aware of the potential risk of conjunctivitis, blepharitis, and rosacea with dupilumab and tralokinumab use. Given the higher risk of AD with IL-17 inhibitors compared to other immunomodulatory medications analyzed in this study, IL-17 inhibitors should be used cautiously in PsO patient populations who are more prone towards developing AD, such as those with a prior history of atopy(*29*). Class switching to therapeutics more likely to prevent a phenotypic switch towards IL-22, such as the anti-IL-23s, or using JAK inhibitors to block upstream of IL-22, might be effective ways of treating emergent AD due to anti-IL-17 therapy. New therapies, such as the IL-22 inhibitor fezakinumab(*78*), or the IL-22 receptor blocker temtokibart, may be future options(*79*).

Our study adds further evidence to the immunological heterogeneity of atopic dermatitis and psoriasis(*11*, *80*). Rather than being only a T_H_2 or only a T_H_17 condition, our study suggests multiple immune phenotypes for both conditions exist. These immune phenotypes can switch into another through phenotype fluidity. We believe this is because the emergence and presentation of immune-mediated conditions are not dependent on the frank existence or the absolute values of a particular immune subtype, but instead on the relative ratios between different immune cell populations and their interactions with an individual’s genetics and environment.

There are several limitations to this study. Information in FAERS has not been verified, and thus causation cannot be established. Although signal detection can show the likelihood of an association occurring, it cannot definitively conclude whether an adverse event is the direct result of medication use. The information has not been verified, and collider bias can exist within the data. Case reports of the AD-PsO phenotypic switch in the literature can also lead to over or under-signaling in FAERS for different immunomodulatory therapies due to selection bias(*38*, *81*). Given the limitations, this study is only hypothesis-generating. We encourage further research into understanding the pathogenesis of the AD-PsO phenotypic switch.

## MATERIALS AND METHODS

### Study Design

This study is a retrospective observational analysis using safety surveillance data from the FDA Adverse Events Reporting System from Q1 2015 to Q3 2024. Only therapies approved for at least one year were included in the analysis. Duplicate reports were removed.

### The FDA Adverse Events Reporting System

The FDA Adverse Events Reporting System is a publicly available database containing adverse event reports, medication error reports, and product quality complaints submitted to the FDA by consumers, healthcare providers, and other members of the public (https://www.fda.gov/drugs/surveillance/fdas-adverse-event-reporting-system-faers). The database supports the FDA’s post-marketing safety surveillance for drugs and therapeutic biologic products. The information adheres to the international safety reporting guidance issued by the International Conference on Harmonization (https://ich.org/page/e2br3-individual-case-safety-report-icsr-specification-and-related-files) and is coded according to the Medical Dictionary for Regulatory Activities (MedDRA) terminology (https://www.fda.gov/drugs/cder-international-program/international-regulatory-harmonization). To ensure there was enough data, only therapies that have been approved by the FDA for use in the United States at least one year prior to the date of the last quarterly data analyzed were included in the analysis.

### Initial data processing and cleaning

Raw data was imported from FAERS into Google Colab Pro (https://colab.research.google.com) from Q1 2015 to Q3 2024. After extracting the zip files, we merged all the individual quarterly demographics (DEMO) files together, which contained patient demographic and administrative information in a single record for each event report. This was also done for files containing the drug information (DRUG), the MedDRA-coded reaction (adverse event) files (REAC), and the MedDRA-coded indication for use (INDI) for each individual case report. These four FAERS data sub-sets were the American Standard Code for Information Exchange files used in the analysis. To remove case duplicates, we took the most recent report based on unique case identifications and the latest date the FDA received date from the demographics data and counted the total number of cases after deduplication.

### Dupilumab and tralokinumab analysis

To find the values needed for the disproportionality analysis for dupilumab and tralokinumab, the DRUG data was searched for all rows containing dupilumab or tralokinumab in the product active ingredient column. This filtered column was combined with the deduplicated DEMO data to make sure every case was unique, and then it was combined with the REAC data.

The new dataset was imported into R 4.3.2 on R studio and the total number of adverse events for each drug was counted, after merging biosimilars (such as tralokinumab-ldrm) with their original biologic. To find the adverse events of interest, the DEMO and REAC datasets were merged, and the preferred term (PT) adverse event column was searched for the following keywords: psoriasis, psoriasiform, arthritis, ankylosing, colitis, crohn, diabetes, lupus, sclerosis, conjunctivitis, blepharitis, and rosacea. This data was then combined with the DRUG dataset and imported into R, where the total number of adverse events of interest were then counted to use in the disproportionality analysis.

The dataset was filtered for cases where the product active ingredient was dupilumab or tralokinumab to find the number of occurrences for the adverse events of interest related to dupilumab or tralokinumab use.

To generate the data where atopic dermatitis, eczema, or dermatitis was an indication for use, the deduplicated DEMO/REAC data, after being filtered for the aforementioned keywords, was combined with the INDI dataset. The combined dataset was then filtered for atopic dermatitis, eczema, or dermatitis in the preferred terminology indications column (INDI_PT). This data was then combined with the DRUG dataset and imported into R to be filtered for dupilumab and tralokinumab. The values for each of the adverse events of interest was then recorded for use in the disproportionality analysis. The deduplicated DEMO/REAC data was also separately combined with the DRUG dataset and filtered for atopic dermatitis, eczema, or dermatitis in the preferred terminology indications column to look for the total number of adverse events associated with dupilumab or tralokinumab when they were used for the treatment of atopic dermatitis, eczema, or dermatitis.

### Psoriasis to atopic dermatitis phenotypic switch with psoriasis and inflammatory bowel disease-associated therapies

To calculate the values needed for the disproportionality analysis for the psoriasis to atopic dermatitis phenotypic switch, and to explore inflammatory bowel disease as well, the DEMO/REAC dataset was then filtered in Google Colab Pro for any cases containing the keywords dermatitis or eczema in the PT column. The cases were then counted to find the total number for each adverse event containing the words dermatitis or eczema using MedDRA terminology.

TNF-α inhibitors used for psoriasis include adalimumab, certolizumab pegol, etanercept, golimumab, and infliximab. IL-17 inhibitors include bimekizumab, brodalumab, ixekizumab, and secukinumab. The IL-23 inhibitors include guselkumab, risankizumab, and tildrakizumab. The IL-12/23 inhibitor ustekinumab is used as well. Apremilast, a phosphodiesterase-4 inhibitor is a small molecule disease-modifying anti-rheumatic drug used in psoriasis treatment that was included in this analysis. The JAK-inhibitors Deucravacitinib, ruxolitinib, tofacitinib, and upadacitinib were also included as a control group for psoriasis.

For the inflammatory bowel diseases, the TNF-α inhibitors adalimumab, etanercept, golimumab, and infliximab, the IL-23 inhibitors guselkumab, mirikizumab, risankizumab, and tildrakizumab, and the IL-12/23 inhibitor ustekinumab were analyzed. The JAK-inhibitors tofacitinib and upadacitinib, and the α4β7 integrin inhibitor vedolizumab were explored as well.

To find the total number of adverse events for each immunomodulatory therapy of interest, the DRUG dataset was filtered for adalimumab, certolizumab pegol, etanercept, golimumab, infliximab, bimekizumab, brodalumab, ixekizumab, secukinumab, guselkumab, risankizumab, tildrakizumab, ustekinumab, apremilast, deucravacitinib, ruxolitinib, tofacitinib, and upadacitinib, along with the inflammatory bowel disease medications mirikizumab and vedolizumab in Google Colab. The filtered data was then merged with the deduplicated DEMO data and imported into R to get counts for the total number of adverse events for each medication, after cleaning the data by merging biosimilars to their parent biologic. The medications for each class were then added up to get the class-wide numbers of adverse events if the medication belonged to a class.

The dataset containing only dermatitis and eczema reactions was then imported into R. After merging biosimilars with their original biologic, the total number of MedDRA-coded adverse events containing either dermatitis or eczema was then counted for each individual systemic therapy and the various classes for later use in the disproportionality analysis.

To generate the values used in the disproportionality analysis for therapies when the indications for use were psoriasis, Crohn’s disease, or ulcerative colitis, the deduplicated DEMO dataset was combined with the INDI dataset that was filtered to only contain rows where the INDI_PT was psoriasis, crohn, or ulcerative on Google Colab. The data was merged with the DRUG data after filtering it for the therapies of interest, mentioned above. The data was filtered with the dataset containing only dermatitis and eczema reactions to keep the rows of interest. This combined dataset was then imported into R and cleaned by merging all biosimilars with their original biologic, allowing us to generate the total number of adverse events for each MedDRA term containing dermatitis or eczema for the medications of interest. The adverse event counts for each biologic were also combined with the same adverse event counts for other biologics in their respective classes to get the class totals.

### Other investigative values

To calculate the values needed for the analysis associated with allergic conditions, and for the occurrence of new cases of inflammatory bowel disease, namely Crohn’s disease and ulcerative colitis, after using medications in the treatment of psoriasis, a process similar to the previous two processes was followed. The keywords used were asthma, allergic, eosinophilic, and rhinitis for allergic conditions, along with crohn and ulcerative for inflammatory bowel disease. The disproportionality analysis was done only for RORs where the indication was psoriasis because we were interested in seeing if a paradoxical formation of allergic and inflammatory bowel conditions exists in the context of psoriasis treatment.

### Disproportionality analysis

The basis of pharmacovigilance research is the generation of adverse event signals through a disproportionality analysis from individual case safety reports(*82*, *83*). Disproportionality in our case refers to the over- or underrepresentation of an adverse event occurring for therapies of interest in comparison to all other combinations of drugs and events(*84*).

|  |  |  |  |
| --- | --- | --- | --- |
|  | Exposure to<br>Drug of interest | Exposure to<br>other Drugs | Sums |
| Occurrence of Adverse Event of<br>interest | DE | dE | E |
| Occurrence of other Adverse<br>Events | De | de | e |
| Sums | D | d | N |

Where the capital D refers to the use of the drug of interest, lowercase d referring to the use of all other drugs excluding the drug of interest, capital E referring to the occurrence of the adverse event of interest, and lowercase e referring to the occurrence of all other adverse events. Thus, DE refers to the occurrence of the adverse event of interest after exposure to the of the drug of interest, De referring to the occurrence of other adverse events after exposure to the drug of interest, dE referring to the occurrence of the adverse event of interest after exposure to other drugs, and de referring to the occurrence of all other adverse events after exposure to all other drugs. N refers to the total number of reports(*84*).

The reporting odds ratio is one method used to calculate disproportionality. It is the odds ratio used in pharmacovigilance studies. It looks at the odds of a drug and adverse event combination (DE) occurring over the total number of other adverse event for the drug of interest (De) divided by the odds of a given adverse event occurring for all other drugs excluding the drug of interest (dE) over the occurrence of other adverse events for all other drugs (de).

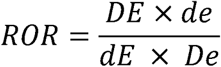

The ROR is positively skewed but approximately has a normal distribution after z-transformation with log or natural log (ln)(*84*). To find the confidence interval, the ROR was converted to ln, applied 1.96 times the standard deviation, and was converted back to its original scale to find the confidence interval with p ≤ 0.05(*84*).

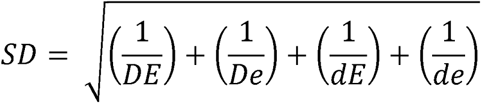

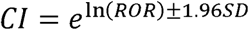

To consider a signal as statistically significant, the lower boundary of the ROR confidence interval must be greater or equal to 1 (ROR_lower_ ³ 1). value of 3.84 for p ≤ 0.05 given 1 degree of freedom(*84*).

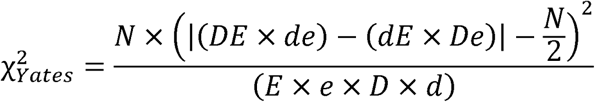

A chi-squared with Yates’ correction was calculated to prove non-normal distribution at a cutoff Given the risk of random incidental reporting with adverse event databases, a minimum number of 4 occurrences is required for a signal to be considered positive(*85*).

The DE, D, E, and N values from FAERS were obtained from the data cleaning and processing on Google Colab and R as described previously. The other values were calculated as follows:

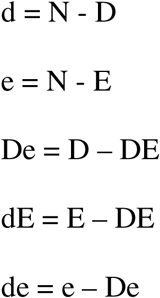

From these values, the ROR, confidence intervals, and χ^2^ were calculated to determine whether a signal is considered statistically significant.

### Change in eczematous dermatitis signal strength

Adverse events classified under the MedDRA codes of allergic dermatitis, atopic dermatitis, contact dermatitis, dermatitis, dyshidrotic dermatitis, eczema, eyelid eczema, hand dermatitis, neurodermatitis, nummular eczema, psoriasiform dermatitis, seborrhoeic dermatitis, and stasis dermatitis were considered to be eczematous dermatoses for the purpose of the study.

Selection bias can exist within pharmacovigilance studies due to a reporter having knowledge that a studied factor is known to cause or is likely to cause the event of interest. This can cause better known adverse events to be overreported and lesser-known adverse events to be underreported. To minimize this bias, each eczematous dermatitis was given equal weighting, and the average ROR was computed on GraphPad Prism for TNF-α, IL-17, IL-23, IL-12/23, apremilast, and JAK inhibitors in general, and when the treatment indication was set to psoriasis or the inflammatory bowel diseases, if the medications were used in the treatment of inflammatory bowel diseases. The average RORs for each medication or class were compared when the treatment indications were narrowed.

Figures 1 to 5 were created on GraphPad Prism. Figure 6 was created in BioRender.

## Data Availability

All data produced in the present study are available upon reasonable request to the authors

## Acknowledgments

We would like to thank S. Nyandwi for his helpful contributions to this project.

## Funding

The authors received no specific funding for this work.

## Author contributions

Conceptualization: KY, CGB, FJ Methodology: KY, AM, CGB, FJ Investigation: KY, IV, GD, CGB, FJ Visualization: KY, AM, DO, SM, CZ, CGB, FJ Project administration: IV, GD, CGB, FJ Supervision: CGB, FJ

Writing – original draft: KY, AM, DO, CGB, FJ

Writing – review & editing: KY, AM, DO, SM, IV, GD, CGB, FJ

## Competing interests

Authors declare that they have no competing interests.

## Data and materials availability

All data, code, and materials are available upon request to the corresponding authors.

## Notes

### Competing Interest Statement

The authors have declared no competing interest.

### Funding Statement

This study did not receive any funding

